# Intestinal parasitic infections associated with nutritional status and inflammatory markers among young children in Huye district, Rwanda

**DOI:** 10.1101/2024.10.07.24315053

**Authors:** Wellars Twahirwa, Xavier Nyandwi, Jean D’Amour Iradukunda, Jean Felix Muneza, Philbert Kanimba, Khadijat O. Adefaye, Noel Gahamanyi, Nadine Rujeni

## Abstract

**Background:** Intestinal parasitic infections (IPIs) are a public health issue affecting young children in low and middle income countries (LMICs). These factors may induce malnutrition, as well as systemic and/or intestinal inflammation, depending on the species, intensity of infection, and host response. This study aimed at determining the effect of intestinal parasites on nutritional status and inflammatory responses in pre- and school-aged children in rural areas of the southern province of Rwanda.

**Methods:** A cross-sectional study involving 127 children under 12 years of age was conducted at two health centers in Huye District, Southern Province, Rwanda, from January to February 2022. A structured questionnaire was used to collect sociodemographic information, feeding habits, anthropometric measurements, and information on infection/malnutrition risk factors. Stool samples were collected to test for intestinal parasites by using microscope, while serum was collected to measure (anti)inflammatory markers [interleukin-10 (IL-10), tumour necrosis factor-alpha (TNF-α), total protein, and C-reactive protein (CRP)].

**Results:** The overall prevalence of IPIs was 38.6%, with non-pathogenic *Entamoeba coli* being the most prevalent (21.3%), followed by *Ascaris lumbricoides* (18.1%), *Entamoeba histolytica* (11.8%), and *Trichuris trichiura* (1.6%). Coinfections accounted for 12.6% of the infections. Moreover, 48.0%, 25.2%, and 9.4% of the children were stunted, underweight, and stunted, respectively. Underweight, IL-10, and total protein levels were significantly associated with IPIs. Our findings also indicated that food supplements had a significant positive effect on stunting.

**Conclusion:** *Ascaris lumbricoides*, *Entamoeba histolytica*, and *Trichuris trichiura* were the predominant parasites. Intestinal parasitic infections in preschool children and schoolchildren affect the nutritional status, possibly through chronic inflammation. Further mechanistic investigations will shed more light on the regulation of the inflammatory response.

## INTRODUCTION

Human intestinal parasites are grouped into protozoan and intestinal helminths, which cause intestinal parasitic infections (IPIs) in the gastrointestinal tract (1). These infections may induce systemic and/or intestinal inflammation, depending on the species, intensity of infection, and host response. Globally, Soil-transmitted helminths infect more than 2.5 billion people worldwide, while protozoa infect half a billion people worldwide (2,3). These parasites are mainly transmitted via the fecal-oral route, although some can be acquired through skin penetration. In 2021, the World Health Organization (WHO) reported that 149.2 million young children under the age of five were stunted, 45.4 million were wasted, and 38.9 million were overweight (4).

Environmental enablers of intestinal parasites include warmth and humidity, which favour their life cycle (5). Moreover, these parasites are associated with poverty, malnutrition, crowded populations, improper fecal disposal, poor sanitation, unclean water, and a low level of education (5,6). In low and middle income countries (LMICs), there is a high prevalence of IPIs, and while they affect all groups of people, pre- and school-aged children carry the heaviest burden (3,7–9).

The immature immune system of children and exposure to poor hygienic conditions increase susceptibility to IPIs (6). IPIs can lead to and/or exacerbate malnutrition via damage to the intestinal mucosa and interference with nutrient absorption (10–12), while malnutrition can increase susceptibility to IPIs (13). These parasitic infections can negatively contribute to the loss of various nutrients and affect nutritional status (6,9). A deficiency of some micronutrients can reduce the body’s immune function and increase the susceptibility of the host to infections (13–16). Malnutrition and IPIs are believed to impair the immune system (17–19). Chronic inflammation in children is a leading cause of mortality, morbidity, language development difficulties, late growth, mental health conditions, poor school performance, and stunting (10,14,15). A number of studies have suggested that inflammation due to helminth infections may indirectly affect the central nervous system (CNS) and negatively affect neurodevelopment in children (20,21).

Soil-transmitted helminths (STHs) promote systemic inflammation by inducing anti-inflammatory cytokines such as IL-10, which are antagonistic to proinflammatory cytokines such as TNF-α, IL-6 and leptin, which are secreted by adipose tissues (22). Infections established by STHs are characterized by a T-helper cell type 2 (Th2) response, whereas regulatory cytokines (e.g., IL-10 and TGFβ) are induced to limit the effect of these responses on helminths and protect the host from immunopathology (23–25). Furthermore, helminths polarize macrophages to an alternatively activated phenotype characterized by Arg-1 and nitric oxide synthase 2 (NOS2) upregulation, downregulation of proinflammatory factors such as IL-6 and TNF-α, and secretion of IL-4, IL-13, and IL-5 in macrophages, innate lymphoid cells 2 (ILC2s), and eosinophils (21). The role of the proinflammatory cytokine TNF-α in gastrointestinal helminths is to induce the Th2 response (26).

In Rwanda, a high prevalence of IPIs has been recorded in various parts of the country (27–29). On the other hand, various efforts have been made by the Government of Rwanda to improve the nutritional status of children by providing food supplements (*Shisha kibondo*) to children from poor families, training parents on how to prepare a balanced diet for children, building kitchen garden*s,* one cow per family *(Girinka munyarwanda)*, early childhood development centers (*Irerero)*, and deworming campaigns. In 2020, the Rwanda National Child Development Agency (NCDA) reported that the national prevalence of malnutrition decreased from 38% in 2015 to 33% in 2020 in children under five years of age (30).

However, no studies have been conducted to explain the relationship between intestinal parasites infections, nutritional status and inflammation among children in Rwanda. Therefore, the aim of this study was to assess the effect of intestinal parasitic infections on nutritional status and inflammation among pre- and school-aged children from rural areas of Huye District, southern province of Rwanda.

## METHODS

### Study design

A cross-sectional study was conducted at the Sovu and Rango Health Centers (HCs) in HUYE District, Southern province of Rwanda, from January to February 2022; the study was carried out on children aged 2-12 years attending Sovu and Rango HCs. A total of 127 children were recruited and assessed for parasitological parameters, Z scores, and risk factors. The number was further reduced to 80 when it comes to the analysis of inflammatory cytokines.

### Ethical consideration

The study was approved by the CMHS Institutional Review Board (ethical clearance no CMHS/IRB/355/2021) and KABUTARE District Hospital. In this study, the parents or guardians of the participants were informed of the aim of the study and asked to sign a consent form on behalf of their children. Participation was voluntary, and participants had the right to withdraw at any stage of the study.

### The eligibility criteria

All children aged 2-12 years attending Rango and Sovu Health Centers from 7^th^ January to 12^th^ February 12, 2022 whose parents or guardians agreed to participate were included in this study. Those previously treated for intestinal parasites for at least one month were excluded.

### Data collection method and examination

#### Questionnaire and consent form

A structured questionnaire was used to collect information from each participant. It was developed based on known associated risk factors and it was first prepared in English and then translated into the Kinyarwanda (local language). The aim of the study was explained to the parents before collecting blood and stool samples and they were asked to sign a consent form.

#### Anthropometric measurements

The measurements included weight, length but we also recorded the age of the children. The nutritional status of the child (weight for age, height for age, and body mass index (BMI) for age) was also determined. The height of each child was measured using a length board and recorded in centimeters (cm) whereas a digital balance scale was used to measure weight and recorded in kilograms (kg). During height measurement, the heels of the child were attached to the wall of the length board, the legs were straight, the arms were on the side, the head looked forward, and the shoulders relaxed (31).

To measure weight, the children who could stand alone were requested to stand independently at the center of the scales, and those who could not stand alone were weighed along with the mother. Subsequently, the weight of the child was calculated by subtracting the weight of the mother alone.

The weight of each child was measured in minimum clothes, without socks and shoes, and was recorded with a precision of 0.1 kg, and the length was measured and recorded in centimeters (cm).

#### Sample collection and storage

Approximately, 4 ml of blood were collected according to World Health Organization (WHO) guidelines (32). After blood collection, tubes were labelled with the participant code and sterile stool containers were labelled with the IDs and given to the parents or children for stool sample collection. Collected stool samples were immediately preserved in 10% formalin. Sera samples were separated from whole blood by centrifugation and stored in a −20 °C freezer at the University Teaching Hospital of Butare (CHUB) laboratory. Preserved stool samples were stored in the Microbiology Laboratory at Huye Campus, College of Medicine and Health Sciences, University of Rwanda.

### Stool and blood examination

#### Stool examination

Stool samples were analysed by using the formalin-ether concentration technique by mixing 1 ml of stool with 10 ml of 10% formalin as described by WHO (33). After performing all the procedures recommended by the WHO, two or three drops of normal saline were mixed with the sediment in a tube. One drop of the mixture was added to the slide, and after applying a cover slip, the slides were observed under a microscope at 10X, and then at 40X objectives. Different protozoan cysts and helminth eggs were observed and recorded.

#### Blood examination

Approximately 35 and 45 blood samples from infected and non-infected participants for IPIs, respectively, were selected to randomly assess inflammatory markers. Human TNF-α and human IL-10 were tested using ELISA. A microplate reader was used to measure the absorbance at 450 nm and the concentrations were calculated in pg/ml. Total protein was measured and expressed in mg/dl using a visual machine, whereas C-reactive protein (CRP) was assessed qualitatively using the CRP agglutination test.

### Data analysis

The data were recorded in Microsoft Excel and imported into the Statistical Package for the Social Sciences (SPSS) version 27 for analysis. The WHO AnthroPlus version 1.0.4 was used to calculate the Z scores for nutritional status weight for age (WAZ), height for age (HAZ), and body mass index for age (BAZ). However, this version was unable to calculate the WAZ scores for children aged over 10 years due to a lack of WHO reference data. Children aged > 10 years may exhibit secondary sexual characteristics that may increase their weight and age (34). Z scores < −2 standard deviations were considered underweight, stunted, and wasting for the WAZ, HAZ and BAZ, respectively (35). Descriptive statistics (frequencies and percentages) were computed. Pearson’s chi-square test and Fisher’s exact test were used to analyse differences using SPSS. Statistical significance was assigned for p values less than 0.05.

## RESULTS

A total of 127 children participated in this study and their sociodemographic characteristics are summarized in Table 1. The majority were female (53.5%) and63% of participants were aged between 2 and 6 years. Only 11% had taken food supplements, 66.1% had a kitchen garden in their family, 31.5% did not wash their hands before eating, and 37.8% had domestic animals in their families. The assessment of nutritional status revealed that stunting and wasting were at 48% and 9.4%, respectively. Among the 110 participants who accessed the WAZ, 25.2% were underweight. The prevalence of parasites was 38.6%.

**Table 1.**
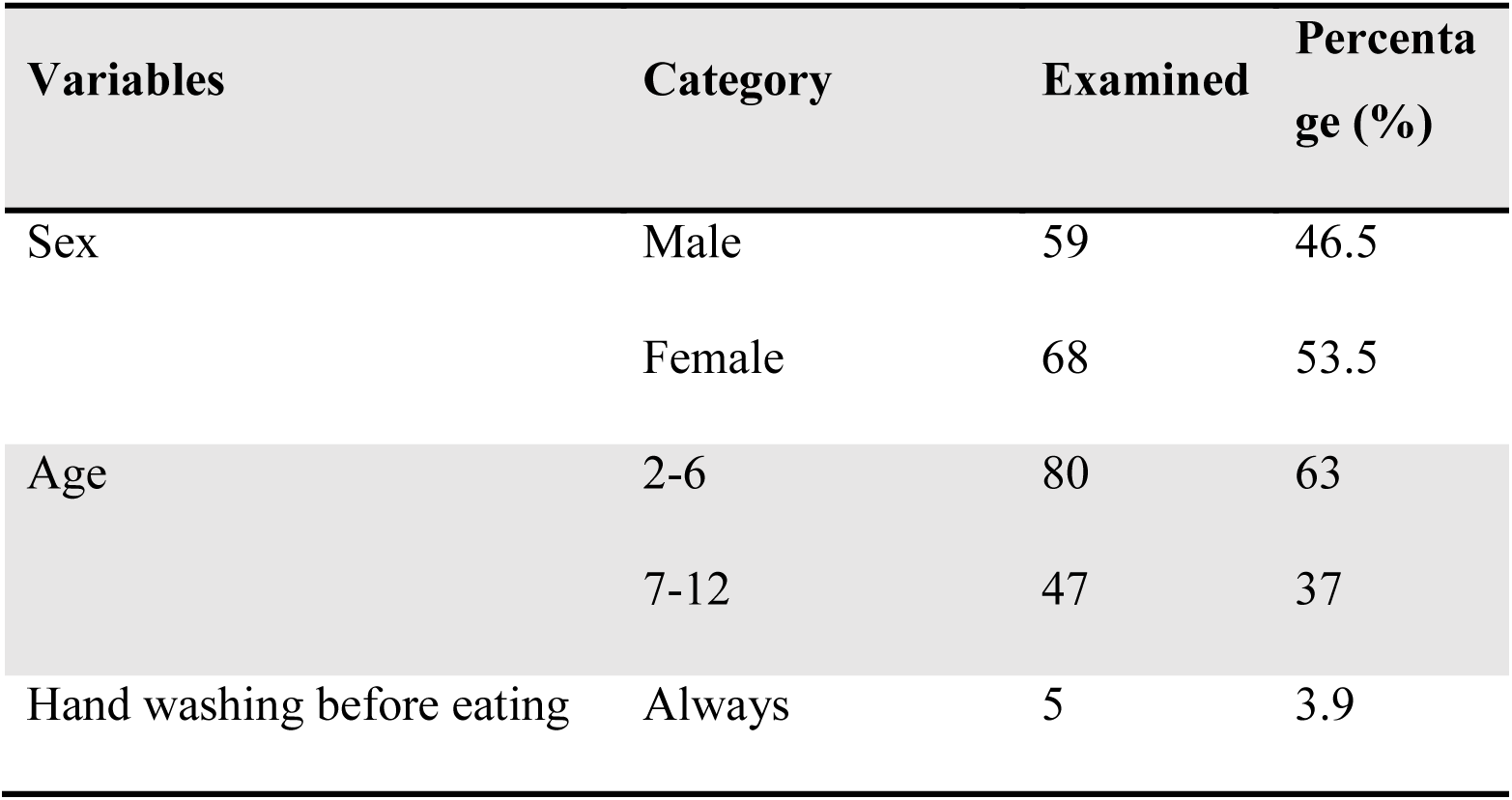
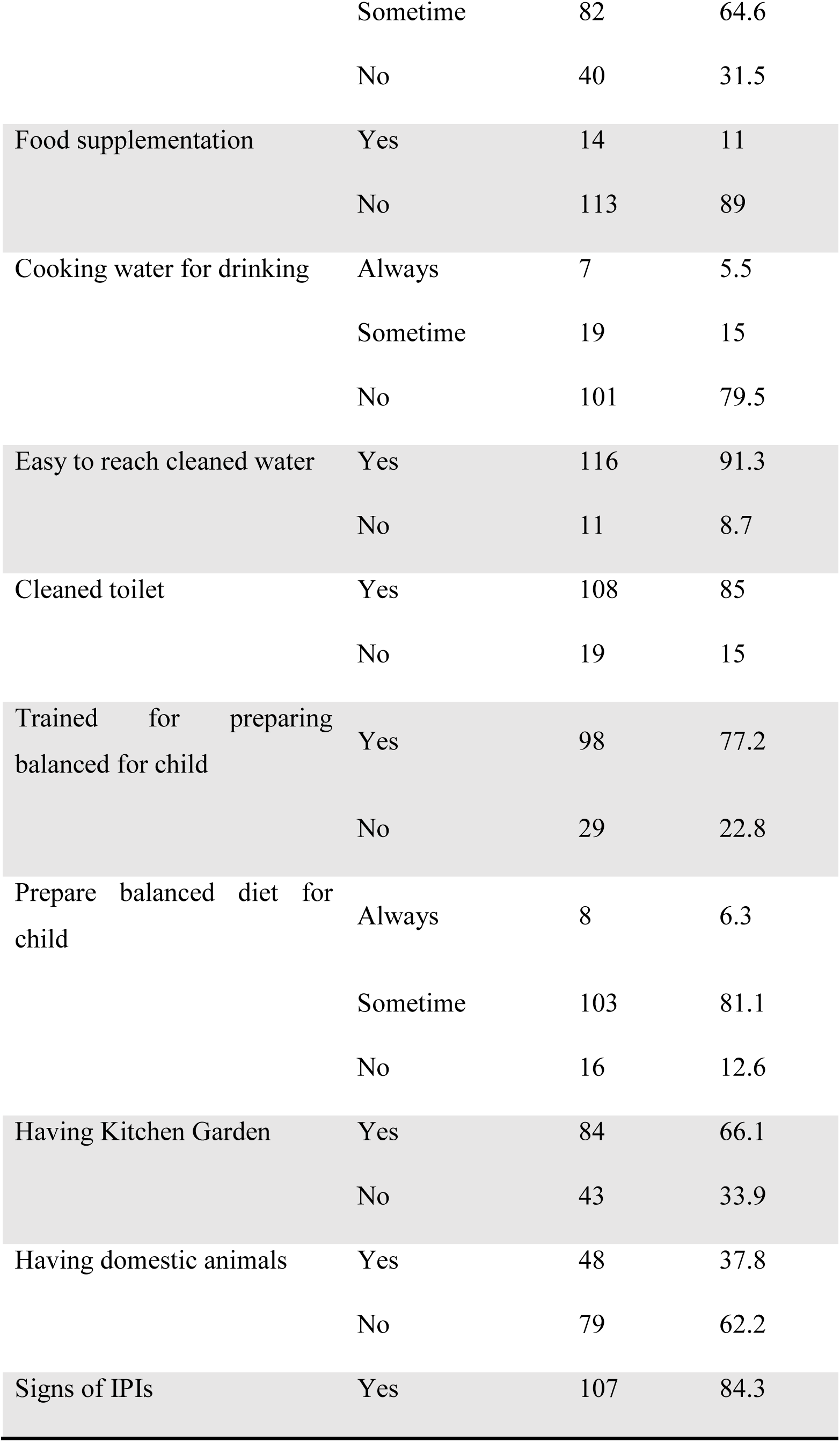
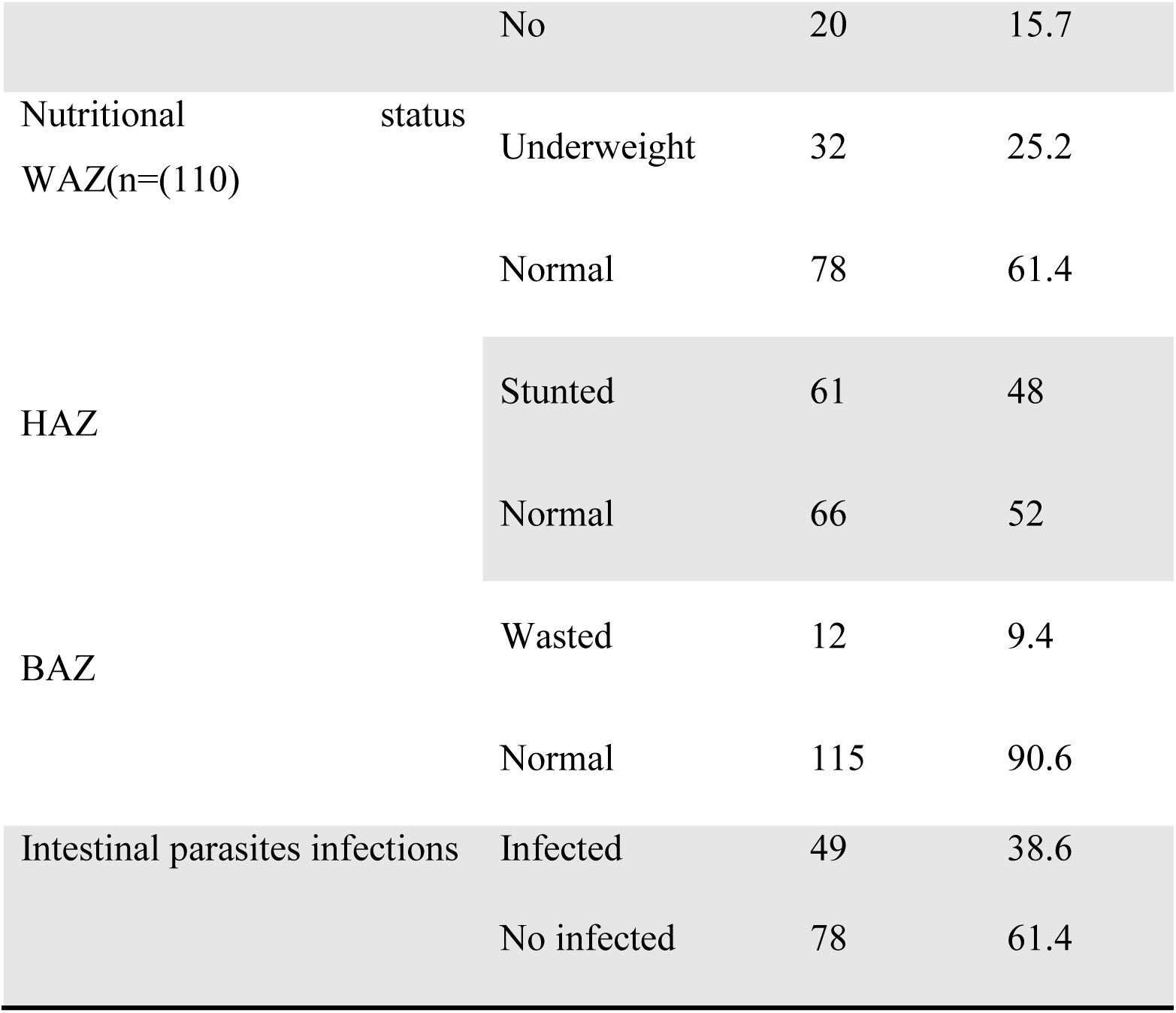
Sociodemographic characteristics of study population (n=127)

### Predominant parasites

The distribution of intestinal parasites in the study population was also assessed. Nonpathogenic *Entamoeba coli* was the most prevalent (21.3%), followed by *Ascaris lumbricoides* (18.1%), *Entamoeba histolytica* (11.8%) and *Trichuris trichiura* (1.6%). Moreover, 12.6% of the children had more than one parasite (coinfection) (Figure 1).

**Figure 1:**
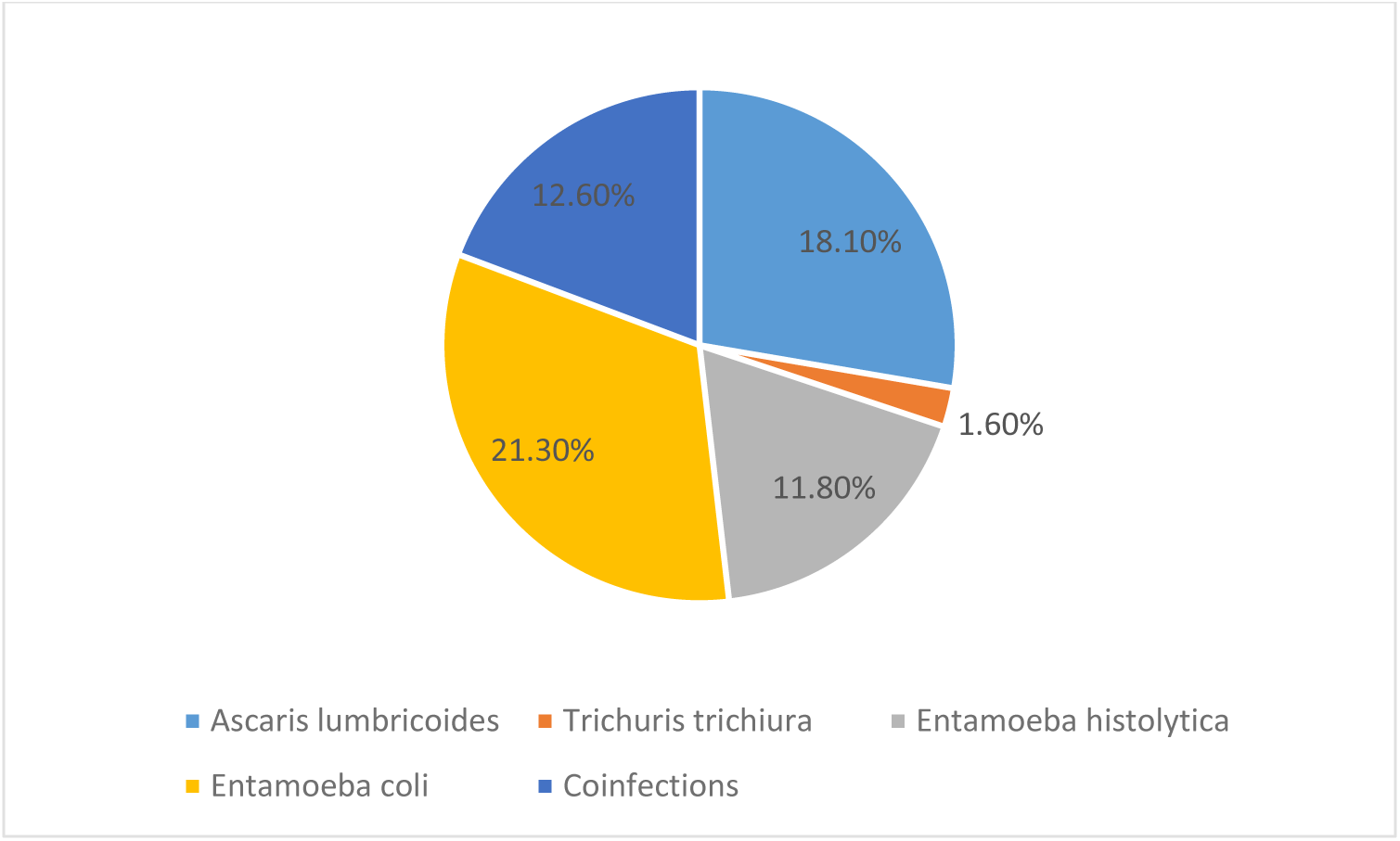
Distribution of intestinal parasites among the study population (n=127)

### Association between IPIs and nutritional status

There was an association between IPIs and the nutritional status of the population. A statistically significant association was seen between IPIs vs underweight (p<0.001), IPIs vs stunting (p=0.002), and IPIs vs wasting (p=0.001) (Table 2)

**Table 2.** Association of IPIs with nutritional status of study population (n=127)

| Variables | Category | Total<br>examined | Not<br>infected<br>No.(%) | Infected<br>No. (%) | X<sup>2</sup> | P value |
| --- | --- | --- | --- | --- | --- | --- |
| WAZ |  |  |  |  |  |  |
| (n=110) | Underweight | 32 | 20 (62.5) | 12(37.5) | 105.679 | <0.001 |
|  | Normal | 78 | 50(64.1) | 28(35.9) |  |  |
| HAZ |  |  |  |  |  |  |
|  | Stunted | 61 | 34(55.7) | 27(44.3) | 110.823 | 0.002 |
|  | Normal | 66 | 44(67.6) | 22(32.4) |  |  |
| BAZ | Wasted | 12 | 5(41.7) | 7(58.3) | 109.768 | **0.001** |
|  | Normal | 115 | 73(63.5) | 42(36.5) |  |  |

### Association between IPIs and inflammatory cytokines

We also assessed association between IPIs and inflammatory cytokines. A statistically significant association (p=0.029) was observed in an infected child with a normal concentration of IL-10 and high total protein (p=0.022), but there was no association with other inflammatory cytokines (Table 3)

**Table 3.**
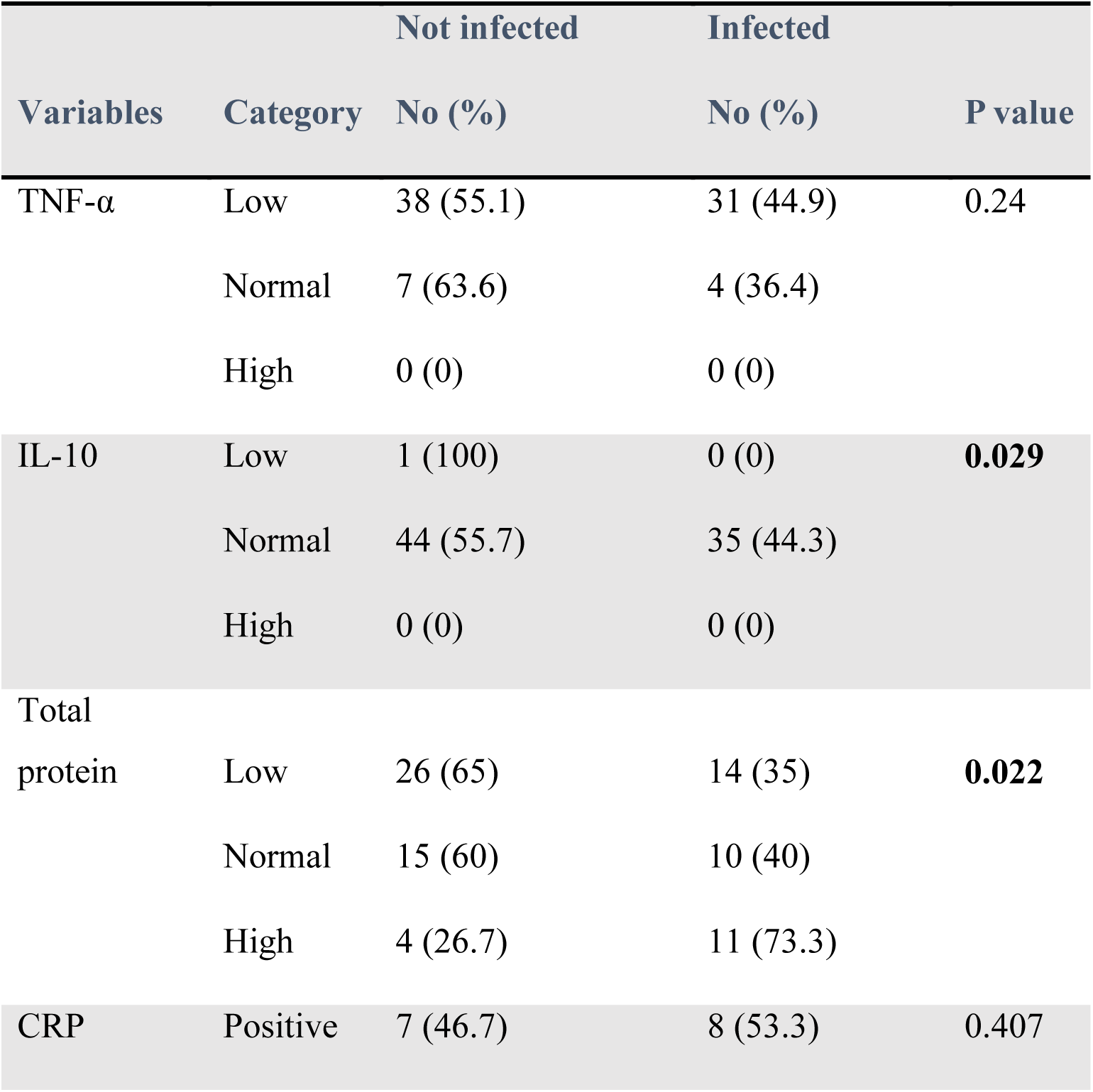
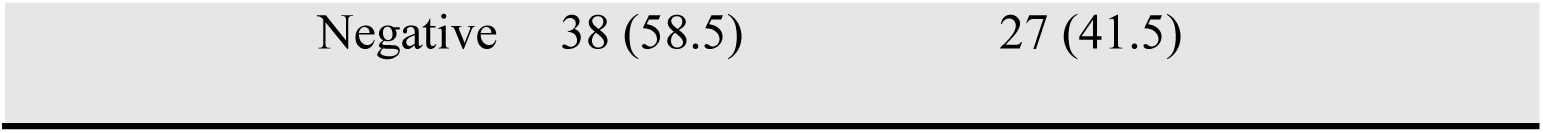
Association of IPIs with inflammatory cytokines (n=80)

|  |  | Not infected | Infected |  |
| --- | --- | --- | --- | --- |
| Variables | Category | No (%) | No (%) | P value |
| TNF-α | Low | 38 (55.1) | 31 (44.9) | 0.24 |
|  | Normal | 7 (63.6) | 4 (36.4) |  |
|  | High | 0 (0) | 0 (0) |  |
| IL-10 | Low | 1 (100) | 0 (0) | 0.029 |
|  | Normal | 44 (55.7) | 35 (44.3) |  |
|  | High | 0 (0) | 0 (0) |  |
| Total |  |  |  |  |
| protein | Low | 26 (65) | 14 (35) | 0.022 |
|  | Normal | 15 (60) | 10 (40) |  |
|  | High | 4 (26.7) | 11 (73.3) |  |
| CRP | Positive | 7 (46.7) | 8 (53.3) | 0.407 |

|  |  |  |
| --- | --- | --- |
| Negative | 38 (58.5) | 27 (41.5) |

### Association between risk factors and nutritional status

We then evaluated the association between promoting risk factors and nutritional status. There was a statistically significant association between food supplement and HAZ (p=0.047). Also, having a kitchen garden was significantly associated with WAZ (p=0.027) and HAZ (p<0.001) but not with other risk factors (Table 4)

**Table 4.** Association between nutritional status and risk factors (n=127)

| Variables | Category | WAZ |  |  | HAZ |  |  | BAZ |  |  |
| --- | --- | --- | --- | --- | --- | --- | --- | --- | --- | --- |
|  |  | Underweight | Normal | P value | Stunted | Normal | P value | Wasted | Normal | P value |
| Food supplement | Yes | 4 | 8 | 0.743 | 3 | 11 | **0.047** | 1 | 13 | 1.000 |
|  | No | 28 | 70 |  | 58 | 55 |  | 11 | 102 |  |
| Kitchen garden | Yes | 16 | 57 | **0.027** | 30 | 54 | **<0.001** | 10 | 74 | 0.336 |
|  | No | 16 | 21 |  | 31 | 12 |  | 2 | 41 |  |
| Trained to prepare balanced diet | Yes | 29 | 62 | 0.266 | 45 | 53 | 0.405 | 10 | 88 | 0.733 |
|  | No | 3 | 16 |  | 16 | 13 |  | 2 | 27 |  |
| Taking balanced diet | Yes | 5 | 3 | 0.128 | 4 | 4 | 0.188 | 0 | 8 | 0.393 |
|  | No | 2 | 6 |  | 11 | 5 |  | 0 | 16 |  |
|  | Sometimes | 25 | 69 |  | 46 | 57 |  | 12 | 91 |  |
| Domestic animals | Yes | 10 | 29 | 0.663 | 27 | 21 | 0.200 | 12 | 115 | 0.764 |
|  | No | 22 | 49 |  | 34 | 45 |  | 7 | 72 |  |

There were associations between intestinal parasites and some risk factors. IPIs were significantly associated with age group (p=0.038), hand washing before eating (p**=**0.038), and cooking water for drinking (p=0.009). However, no associations were observed with other risk factors (Table 5)

**Table 5.**
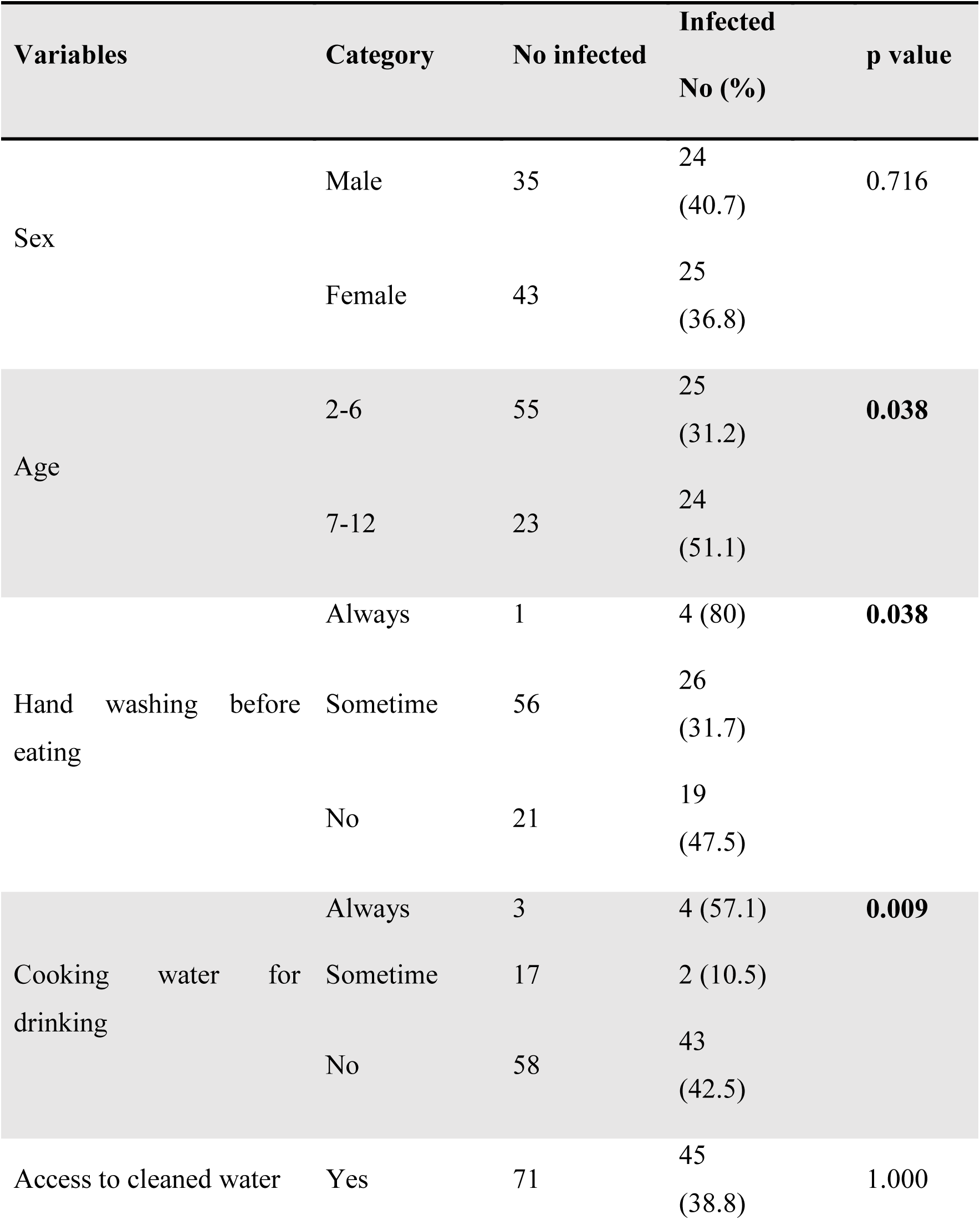
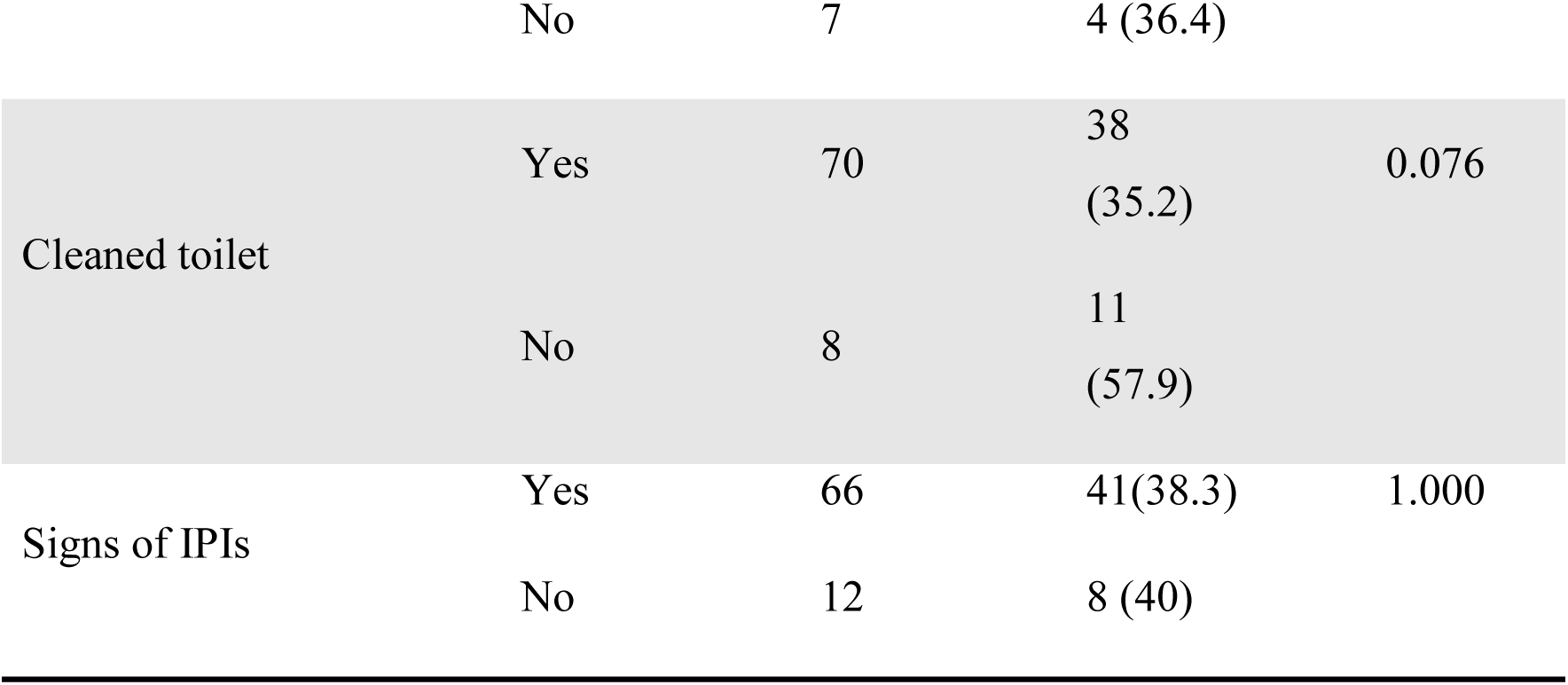
Association of IPIs with risk factors (n=127)

## DISCUSSION

Intestinal parasites and malnutrition cause severe problems in children in LMICs (36). Intestinal parasitic infections have been associated with growth retardation and various health-related problems (15). This study assessed the effect of IPIs on children’s nutritional status and inflammatory response.

The current study reported that the prevalence of IPIs among children living in rural settings in Huye District was 38.6%. The prevalence of this study was lower than 44.8% and 77.7% previously reported in Rwanda (28),(29), 44.3% and 52.9% reported in Ethiopia (37), 34) reported in Ethiopia, 82%and 95.3% previously reported in Pakistan (26, 35). However, the prevalence of this study was higher than 16.6% previously reported in a study conducted in Nigeria (9).The difference may be due to the small sample size of our study, the inability to detect trophozoites of *E. histolytica, E. coli* and *G. lamblia,* which might have been lost during preservation, the season of sampling, and the use of a deworming program in Rwanda.

Older children (aged 7-12 years) had a greater prevalence of infection, which might be due to that the older children in rural areas are engaged in farming activities resulting in exposure to soil and water (6). The same findings were also found in different studies (39–41). In addition, this study revealed a significantly greater prevalence of IPIs among children who ate without washing their hands and who drank unboiled water. The high prevalence of *Entamoeba coli* found in the area, even if considered as non-pathogen, indicates poor hygiene. This is in agreement with previous studies which showed positive association between poor hygiene and IPIs (2,28,42–44).

In this study, the prevalence of stunting, underweight, and wasting were 48.0%, 25.2% and 9.4%, respectively. The high stunting prevalence might be caused by a lack of a balanced diet, especially in those from poor families (3). We found a statistically significant association between IPIs and underweight (p<0.001), stunting (p=0.002), and wasting (p=0.001). These associations are in line with findings from Nigeria (45), Indonesia (5) and Ethiopia (46) that showed a positive association between IPIs and underweight. In contrast, Irisarri-Gutiérrez et al.(27) reported no association between IPIs and nutritional status in the Gakenke District, Northern Province, Rwanda. Children infected with intestinal parasites loose appetite and sometimes nutrients in case of intestinal perforations. This could contribute to status of the children being underweight, stunting, and wasting.

This study revealed an association between IPIs and the systemic anti-inflammatory marker IL-10 (p=0.029) and the marker of malnutrition total protein (p=0.022). This differ from the findings of a study by Zonta et al.(47) conducted in Mexico showing that the increase in IL-10 could be linked to Th2 activation that can increase the predisposition to IPIs and bacterial infections (16). The high total protein concentration observed might have been caused by chronic inflammation resulting from IPIs (13). This study suggested that a high total protein concentration might be associated with intestinal parasite infections. It has been reported that inflammation decreases the total protein concentration by reducing protein synthesis (48). A decrease in the concentration of total protein and body mass index increases the risk of mortality (48), while TNF-α is produced by macrophages and monocytes in the acute phase as a result of the response to parasitic infections. TNF-α increases the production of acute phase proteins such as CRP, ferritin, and complement factors, whereas total protein decreases (13).

Our findings indicate that children who did not consume food supplements had significantly greater levels of stunting, highlighting the positive impact of the program on children’s health in Rwanda. In addition, our study showed a significant association between having a kitchen garden and being underweight and stunted. These findings suggest that low quality and quantity of food intake is a contributor to malnutrition in Huye District. Poverty is the main cause of an inadequate diet, which may affect both the quality and quantity of food, resulting in a lack of nutrients required for the physiological function of the body(20).. No associations were detected between nutritional status and the other associated risk factors in this study. To the best of our knowledge, this is the first study to focus on inflammation in children infected with parasites and with malnutrition in Rwanda.

### Limitations

This study had some limitations. First, it was conducted in rural areas which could differ from urban areas. Second, the sample size was small which could not be representative of the whole district. Larger studies with any observational component will shed light on the possible associations.

## Conclusion

The overall prevalence of IPIs was 38.6% and the predominant parasites were *A. lumbricoides*, *E. histolytica*, and *T. trichiura*. A considerable number of children were stunted, underweight, and underweight. Underweight, IL-10, and total protein levels were significantly associated with IPIs. Improving sanitation and nutrition of children ameliorate the nutritional status of children and their families. Also, regular deworming practices at schools should be encouraged to prevent negative effects of IPIs to the growth and educational outcomes of children

## Acknowledgement

The authors wish to acknowledge all parents/guardians and children who consented to participate in this study. We thank the Institutional Review Board (IRB) of CMHS, KABUTARE District Hospital for approving our study. We acknowledge the guidance of ReachSci Society of the Cambridge University through the ReachSci STEM Mini-PhD program. Lastly, our gratitude goes to the administration of Sovu and Rango Health Centers for supporting this study.

## Authorship contribution

**WT, XN, JDAI**, and **JFM**: Conception, writing review, data analysis, interpretation, and manuscript drafting; **PK:** co-supervision, project administration, writing, and design of the work; **KOA**: interpretation, and manuscript drafting; **NG:** writing original draft, visualisation and, significantly improved the manuscript, and **NR**: supervision, data curation, validation, critical review of the work for important intellectual content, final approval of the version to be published. All authors were involved in writing the paper and provided final approval for publication.

## Data availability statement

All relevant data are within the article.

## Disclaimer

The views and opinions expressed in this article are of those of the authors and do not necessarily represent the official policy or position of any affiliation agency of the authors.

